# Determinants of cardiac adverse events of chloroquine and hydroxychloroquine in 20 years of drug safety surveillance reports

**DOI:** 10.1101/2020.05.19.20107227

**Authors:** Isaac V. Cohen, Tigran Makunts, Talar Moumedjian, Masara Issa, Ruben Abagyan

## Abstract

Chloroquine (CQ) and hydroxychloroquine (HCQ) are on the World Health Organization’s List of Essential Medications for treating non-resistant malaria, rheumatoid arthritis (RA) and systemic lupus erythematosus (SLE). In addition, both drugs are currently used off-label in hospitals worldwide and in numerous clinical trials for the treatment of SARS-CoV-2 infection. However, CQ and HCQ use has been associated with cardiac side effects, which is of concern due to the higher risk of COVID-19 complications in patients with heart related disorders, and increased mortality associated with COVID-19 cardiac complications. In this study we analyzed over thirteen million adverse event reports form the United States Food and Drug Administration Adverse Event Reporting System to confirm and quantify the association of cardiac side effects of CQ and HCQ. Additionally, we identified several confounding factors, including male sex, NSAID coadministration, advanced age, and prior diagnoses contributing to the risk of drug related cardiotoxicity. These findings may help guide therapeutic decision making and ethical trial design for COVID-19 treatment.

## Introduction

Chloroquine (CQ) and hydroxychloroquine (HCQ) were initially used as antimalarial agents with broad-spectrum antiviral effects^1^. The use of CQ worldwide dates back to a 70-year mark with a well-established safety profile as a first line drug for the treatment and prophylaxis of malaria^2^. HCQ, closely structurally related to CQ, has been used at higher doses and for longer duration in some autoimmune diseases such as rheumatoid arthritis (RA) to systemic lupus erythematosus (SLE)^1,2^. Additionally, these agents have been observed and investigated for their antiviral properties^3^. Most common adverse events of (AEs) of these agents include eye disorders, proximal myopathy, neuropathy, neuropsychiatric events, hypoglycemia, worsening of psoriasis, and particularly, cardiac including cardiomyopathy and QT prolongation^4,5^.

Recently, CQ and HCQ were reported as potentially beneficial treatments for severe acute respiratory syndrome coronavirus 2 (SARS-CoV-2) infection, also known as COVID-19, and several clinical studies are currently investigating these drugs for potential therapeutic efficacy^6, 7^ On March 28th, the FDA issued an Emergency Use Authorization of HCQ and CQ products “to be distributed and prescribed by doctors to hospitalized teen and adult patients with COVID-19, as appropriate, when a clinical trial is not available or feasible”^8^. The use of HCQ as an antiviral agent against SARS-CoV-2 was identified in an in vitro study which was a source of encouraging results^9^. Recent trials in China and France reported the potential efficacy of these agents against SARS-CoV-2^6,7^. Specifically, in an early non-randomized controlled trial conducted in France, 100% of the 36 included patients treated with combination HCQ and azithromycin tested negative for SARS-CoV-2 on day 6 post-treatment compared with 12.5% of the controls^6^. Similarly, in a narrative letter, China reported testing CQ and HCQ in patients with COVID-19-induced pneumonia in over 10 hospitals across the nation. The authors had claimed that they have demonstrated the efficacy and safety of CQ phosphate in “more than 100 patients”^7^. Currently, several international ongoing clinical trials are investigating HCQ and CQ for treatment efficacy and prevention (as examples: NCT04315896^10^, NCT04318015^11^, NCT04318444^12^, NCT04321278^13^, NCT04308668^14^, NCT04304053^15^, NCT04316377^16^, and NCT04303299^17^. Despite limited efficacy data, HCQ and CQ are currently being administered as part of first-line treatment for SARS-CoV-2 in various hospitals across the world ^18,19^.

As previously mentioned, cardiac complications attributed to HCQ and CQ have been described in various reports. A systematic review of the literature published in June 2018 examined 86 reports including individual cases and short series from an online database search. The report identified 127 patients with cardiac events attributed to HCQ and CQ administered in the context of various inflammatory disorders with a median daily dose of 250 mg for CQ and 400 mg for HCQ^20^. Recent disproportionality analysis study observed an elevated prevalence of Torsade de Pointes and QT prolongation reports in CQ and HCQ^21^. *The cardiac AEs of these therapeutics are of increased concern since a subset of patients infected with COVID-19 present with cardiac injury, suggesting a relevant cardiovascular involvement in the pathophysiology of the disease^22^*. In a single-centered cohort study from Wuhan, China, Shi et al. examined the incidence of cardiac injury in 416 hospitalized patients with COVID-19. Among those, 82 of the patients demonstrated cardiac injury, and higher in-hospital mortality rates were seen in patients with cardiac issues (51.2%) compared to those without (4.5%)^23^. Guo et al. described similar findings in a retrospective single-center case series analysis in which 52 out of 187 hospitalized patients suffered from myocardial injury. The mortality rate in those patients was 59.6% compared to 8.9% in those without cardiac injury*^24^*. In support of these findings, recent studies have shown that major cardiac outcomes associated with cardiomyopathy (33%) and cardiac injury (23%) are common in critically-ill patients ^25,26^. Similar lines of evidence are also followed by an Italian case report of a 53-year-old woman with lab-confirmed COVID-19 who was admitted to the hospital for severe LV dysfunction and acute myopericarditis. This case highlights that SARS-CoV-2 can impact the cardiovascular system even in the absence of major respiratory tract involvement^27^. Other studies have observed COVID-19 cardiac complications such as fulminant myocarditis, ventricular tachycardia^28,29,30^.

The goal of this study is to reanalyze the extensive clinical data of CQ and HCQ cardiac AEs collected during the last 20 years to derive the strength of the associations and, more importantly, contributing risk factors. The findings may improve the safety of these therapeutics for COVID-19 treatment in patients that are already at higher risk of cardiac complications.

## Methods

### FDA Adverse Event Reporting System

The study used over thirteen million AE reports available from the United States Food and Drug Administration Adverse Event Reporting System (FAERS) and its older version, Adverse Event Reporting System (AERS) data sets. At the time of the study the FAERS/AERS set contained reports from years 2000-2020, all available online at: https://www.fda.gov/drugs/questions-and-answers-fdas-adverse-event-reporting-system-faers/fda-adverse-event-reporting-system-faers-latest-quarterly-data-files

### Data preparation

FAERS/AERS reports are collected through voluntary reporting to the FDA through MedWatch^31^ and stored in quarterly format data subsets with their respective parameters (age, sex, drug, AE etc.), and common case identifiers. FAERS data format has had changes historically, requiring each quarterly set to be individually downloaded and modified into consistent data tables ^32,33,34^. Since the FAERS/AERS set has reports from all over the world with their respective drug brand names, 27 unique terms were recognized and translated into single generic CQ and HCQ names. The final data set contained 13,313,287 AE reports. SLE, RA, and malaria were considered for possible sources of reports, however due to low sample size of malaria treatment with HCQ or CQ in FAERS, only SLE and RA were included in the analysis.

### Cohort selection and data cleaning

702,274 reports were obtained from the FDA FAERS database to form three cohorts for analysis by logistic regression: CQ cohort, HCQ cohort and Control cohort. The control cohort for both drugs was defined by reports with RA and SLE patients where HCQ and CQ were not used (n=639,990). The CQ and HCQ cohorts were defined by reports with RA and SLE indication with CQ (n=1,280) and HSQ (n=65,004) was used in addition to other therapeutics. RStudio (Version 1.2.5033) and R (Version 3.6.3)^35^ were employed for data cleaning and logistic regression modeling. FAERS/AERS data sets historically include a small fraction of duplicate reports. The set was scanned for these entries with the R package “`dplyr`” “`distinct`” function and were removed as appropriate. A summary of the records demographic factors is made available in Table 1. In order to define the list of possible cardiac AEs in this database a table was generated and manually checked for errors by the investigators. For a copy of this table of all AEs considered cardiac related see Supplementary Table S1. Similarly, a list of non-steroidal anti-inflammatory drugs (NSAIDs) was generated for use in our analysis and is made available in Supplementary Table S2. Note that the number of NSAIDs did not include Aspirin, as this was to be modeled separately due to expected divergent effects such as higher cardiovascular risk in the patient demographics. During the data cleaning stage, age was limited to a range of 0 to 125 years. For the purpose of our analysis, only values of “`f`” or “`m`” from FEARS/AERS were analyzed. The R package “`dplyr`” function “`mutate`” and “`str_detect`” were employed for counting the number of NSAIDs and cardiac AEs observed in each report. A subsample of the original database that included reports with non-empty values for age and sex was also prepared. For a summary of the sample size of the subgroups see Supplementary Table S3.

**Table 1.**
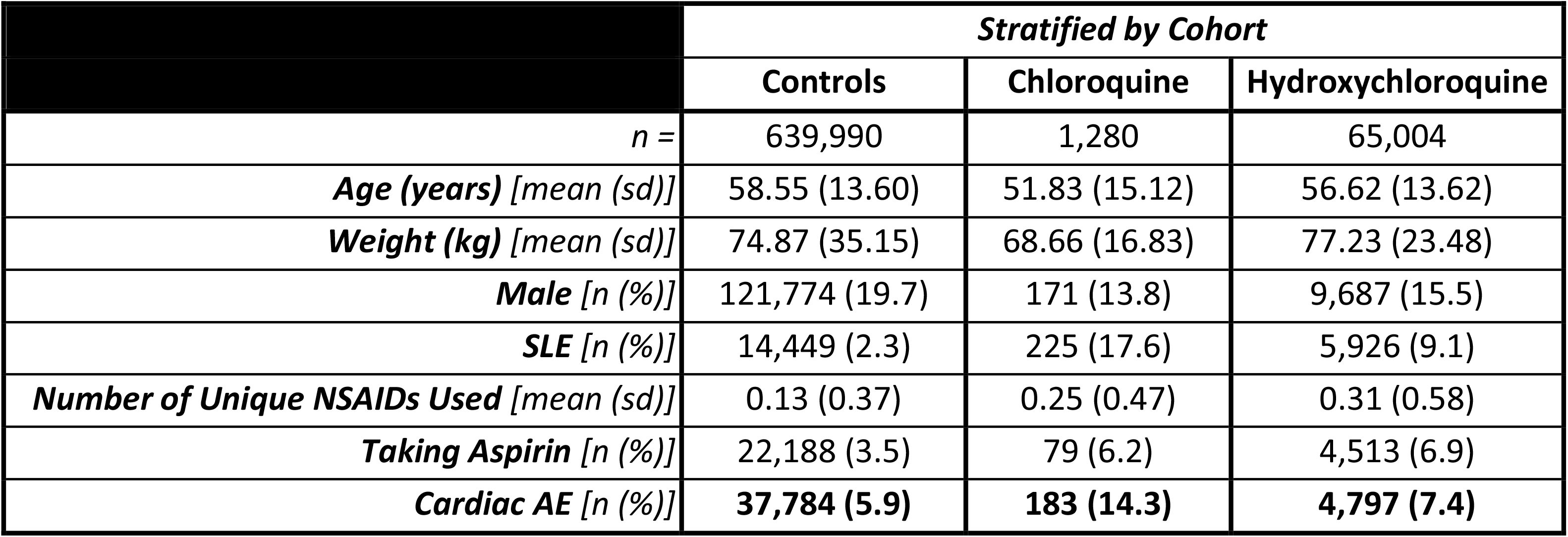
Demographics and distribution of seven selected variables of the three cohorts analyzed by logistic regression. SD is given in parentheses and stands for standard deviation.

|  | *Stratified by Cohort* |  |  |
| --- | --- | --- | --- |
|  | Controls | Chloroquine | Hydroxychloroquine |
| *n* = | 639,990 | 1,280 | 65,004 |
| **Age (years) [mean (sd)]** | 58.55 (13.60) | 51.83 (15.12) | 56.62 (13.62) |
| **Weight (kg) [mean (sd)]** | 74.87 (35.15) | 68.66 (16.83) | 77.23 (23.48) |
| **Male [n (%)]** | 121,774 (19.7) | 171 (13.8) | 9,687 (15.5) |
| **SLE [n (%)]** | 14,449 (2.3) | 225 (17.6) | 5,926 (9.1) |
| **Number of Unique NSAIDs Used [mean (sd)]** | 0.13 (0.37) | 0.25 (0.47) | 0.31 (0.58) |
| **Taking Aspirin [n (%)]** | 22,188 (3.5) | 79 (6.2) | 4,513 (6.9) |
| **Cardiac AE [n (%)]** | **37,784 (5.9)** | **183 (14.3)** | **4,797 (7.4)** |

### Measured outcomes

The primary outcome of interest was incidence of any cardiac AE as defined by one of one of 403 terms listed in Supplementary Table S1. The R package “`gim`” was employed for logistic regression modeling via the “`binomial”` family function. Cardiac AEs were the outcome of interest in logistic regression modeling and were coded as a binary (“`1`” if occurred in the report or “`0`” if not). Covariates explored in the modeling included age (as a continuous variable expressed in years), sex (as a binary), disease state (coded as a binary of either SLE or RA), NSAID usage (as a integer variable equal to the number of NSAIDs observed in the record), and aspirin usage (as a binary). AIC (Akaike Information Criterion)^36^ and n number of records included are reported for each of the eight models presented. Note that due to absence of some values in the raw data from the FDA, models including age and sex have lower sample size than those without these covariates (Supplementary Table S3). Subject weight was not used for model building due to extreme paucity of the data. Models 1a, 1b, 2a, and 2b are built using the control and CQ cohorts (described above and presented in tables 1 and 2, and Supplementary Table S3). Similarly, Models 3a, 3b, 4a, and 4b are built using the control and HCQ cohorts (described above and presented in table 1 and 3, and Supplementary Table S3).F Coefficient estimates, standard error, Adjusted Odds Ratio (Adj. OR), 95% confidence intervals (95%CI), and p-values are reported (tables 2 and 3). P-values that meet the significance threshold of less than 0.05 are marked in tables with an asterisk (*). The adjusted odds ratio is defined as an odds ratio that controls multiple predictor variables in a model and allows for quantification of individual contributions of different variables to a single outcome^37^, in this case cardiac ADRs. The adjusted OR is calculated by the following equation:*_e_^coefficient^*, and is intended to account for biases in association between variables from the sample data.

## Results

### Patient demographic variables

Demographics of the three cohorts to be analyzed for cardiac side effects by logistic regression is presented in Table 1. All three cohorts were comprised of patients who were treated for either SLE or RA. The control cohort (n = 639,990) was compiled from SLE or RA FAERS/AERS reports with no report of CQ or HCQ use. The CQ cohort (n = 1,280) was composed of records of SLE or RA patients with reports of CQ use but not HCQ. Similarly, the HCQ cohort (n = 65,004) was composed of records of SLE or RA patients with reports of HCQ use but not CQ. As shown in Table 1 the mean patient was in their mid-fifties and about 70 kg. The patients were predominantly female, with only 19.7% of the controls listed as male. Additionally, the patients were predominantly treated for RA. The CQ cohort had the most SLE patients at 17.6%. Many of the patients in all three cohorts received various NSAIDs and experienced cardiac adverse events (for a listing of the AEs considered cardiac related see Supplementary Table S1). These numbers of records are large, in particular for HCQ, they cover a range of demographic parameters, and are sufficient to evaluate the contributors to the cardiac AEs with logistic regression. The last row of Table 1 indicates an elevated number of cardiac side effect-containing reports for both CQ and HCQ, from 5.9% to 14.3% and 7.4% respectively. Interestingly, the most common *individual* adverse effects (not grouped by the category) for each cohort were calculated and they did not include cardiac AEs (see Supplementary Fig. S1).

### Chloroquine

Several logistic regression analyses were performed in order to evaluate whether cardiac AEs were related to CQ. Binary logistic regression was employed to determine the confounding variables contributing to the apparent effects of the drugs on occurrence of cardiac side effects. In the first, simplest analysis explored, model 1a (Table 2), CQ was found to significantly increase risk of cardiac AEs (Adj. OR 2.49, p < 2×10^-16^) when controlling for disease state group. Additionally, SLE patients were observed to have higher risk of cardiac AEs (Adj. OR 1.46, p < 2×10^-16^). Model 1b (Table 2) explored NSAID and Aspirin use as possible confounders of these effects. It was found that NSAID and Aspirin use were both predictors of cardiac events, and the effect of CQ on negative outcomes was preserved after accounting for co-administration (Adj. OR 2.24, p < 2×10^-16^). Furthermore, this model’s AIC improved by 6918 compared to model 1 a. Models 2a and 2b (table 2) were built from a subsample of the original database that included records that did not have missing values for age and sex. For a summary of the sample size of the subgroups see Supplementary Table S3. Model 2a shows that sex and gender both effect risk of cardiac adverse events. Male sex was shown to increase risk (Adj. OR 1.57, p 2×10^-16^). Additionally, each year of life was associated with a 2% increased risk of cardiac AEs (p < 2×10^-16^). Model 2b was generated in order to explore effects of NSAID and Aspirin use on the coefficients explored in model 2a. Similar to Model 1b, both aspirin use and NSAIDs were shown to both independently increase risk. Model 2b improved upon model 2a’s AIC by 4934, indicating the significant contribution of the added variables. Moreover, CQ’s deleterious effects on cardiac AE risk in the more complex model 2b were still profound, with an adjusted odds ratio of 2.46 (p < 2×10^-16^).

**Table 2.**
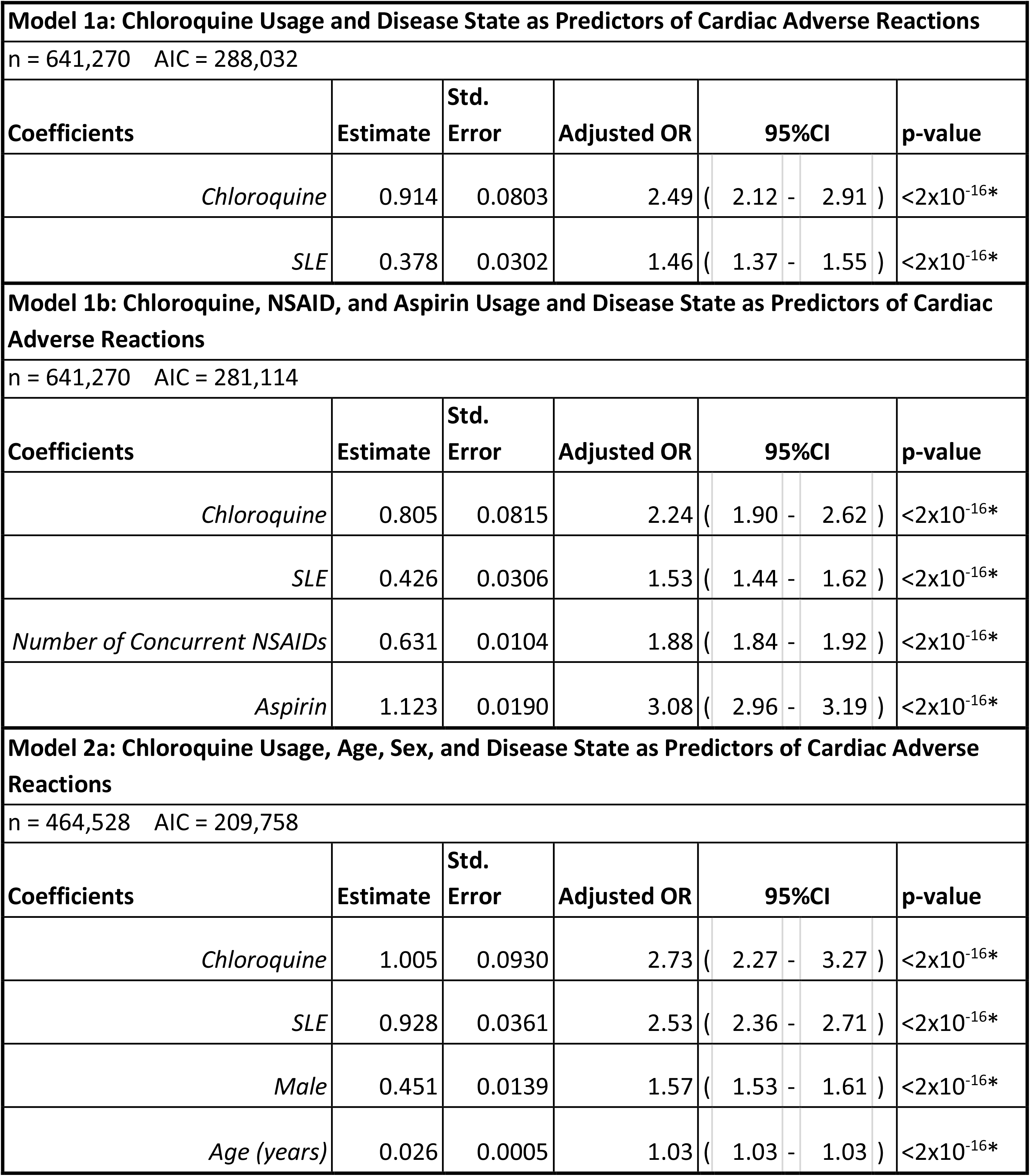
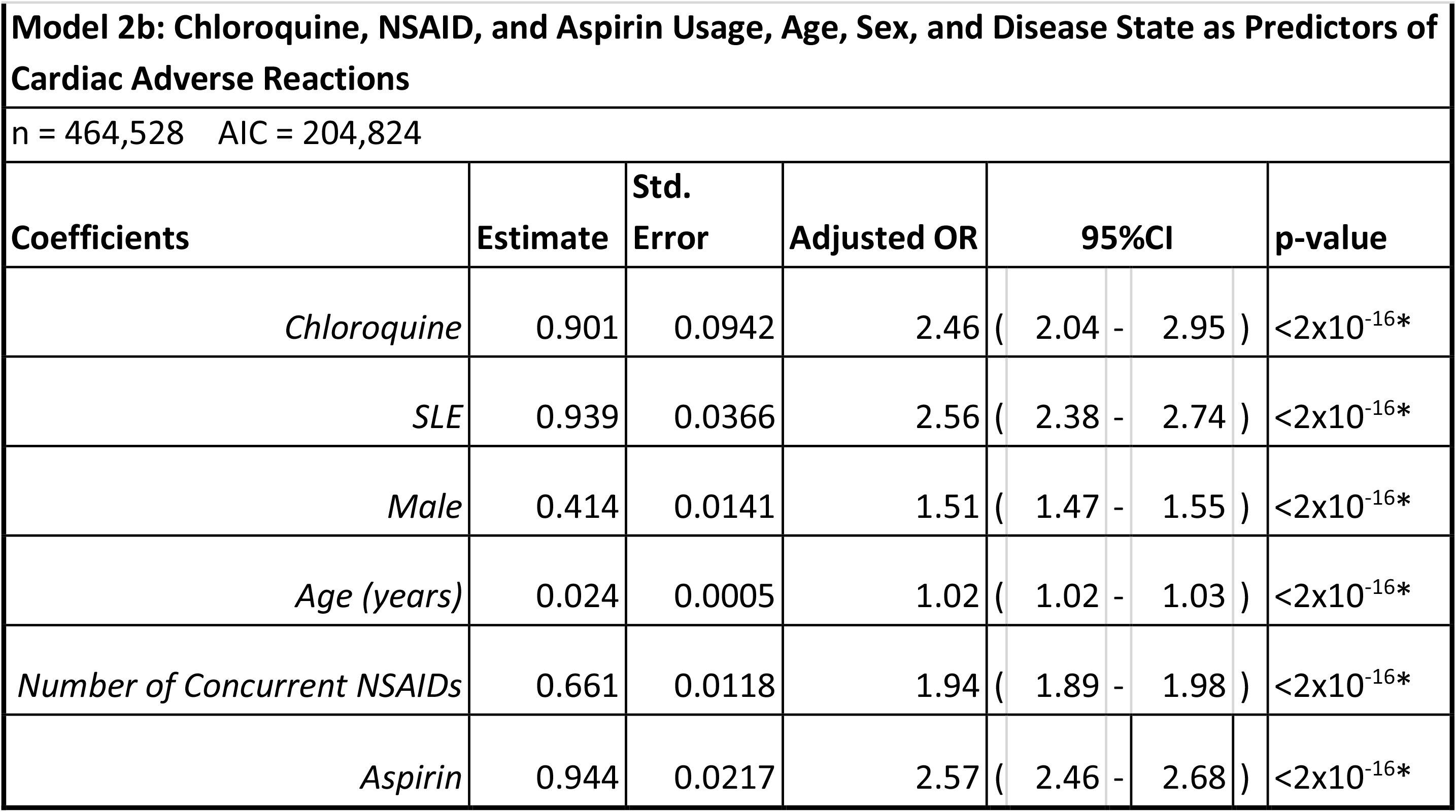
Logistic regression analysis of chloroquine AE reports, * = coefficients significant p-values.

### Hydroxychloroquine

HCQ cardio AE analysis reveals a similar pattern to the results presented in table 2. Table 3 explores these same effects due to HCQ in RA and SLE patients. SLE, NSAID use, aspirin use, male sex, and advanced age were all shown to be important factors in predicting cardiac AEs (Table 3). In the simplest model, model 3a, HCQ was shown to significantly increase cardiac AEs by a modest adjusted odds ratio of 1.22 (p < 2×10^-16^). Interestingly, in model 3b this effect was lost after controlling for NSAID and aspirin use (p = .51). Surprisingly, after adding age and sex into the model this effect’s significance was regained (Model 4b, Adj. OR 1.15 [95%CI 1.10-1.19], p = 8.2×10^-13^). Although modest, HCQ has been shown to have significant effects on patients’ risk of cardiac adverse reactions. This risk has also been shown to be exacerbated by both clinical and demographic factors (Table 3).

**Table 3.**
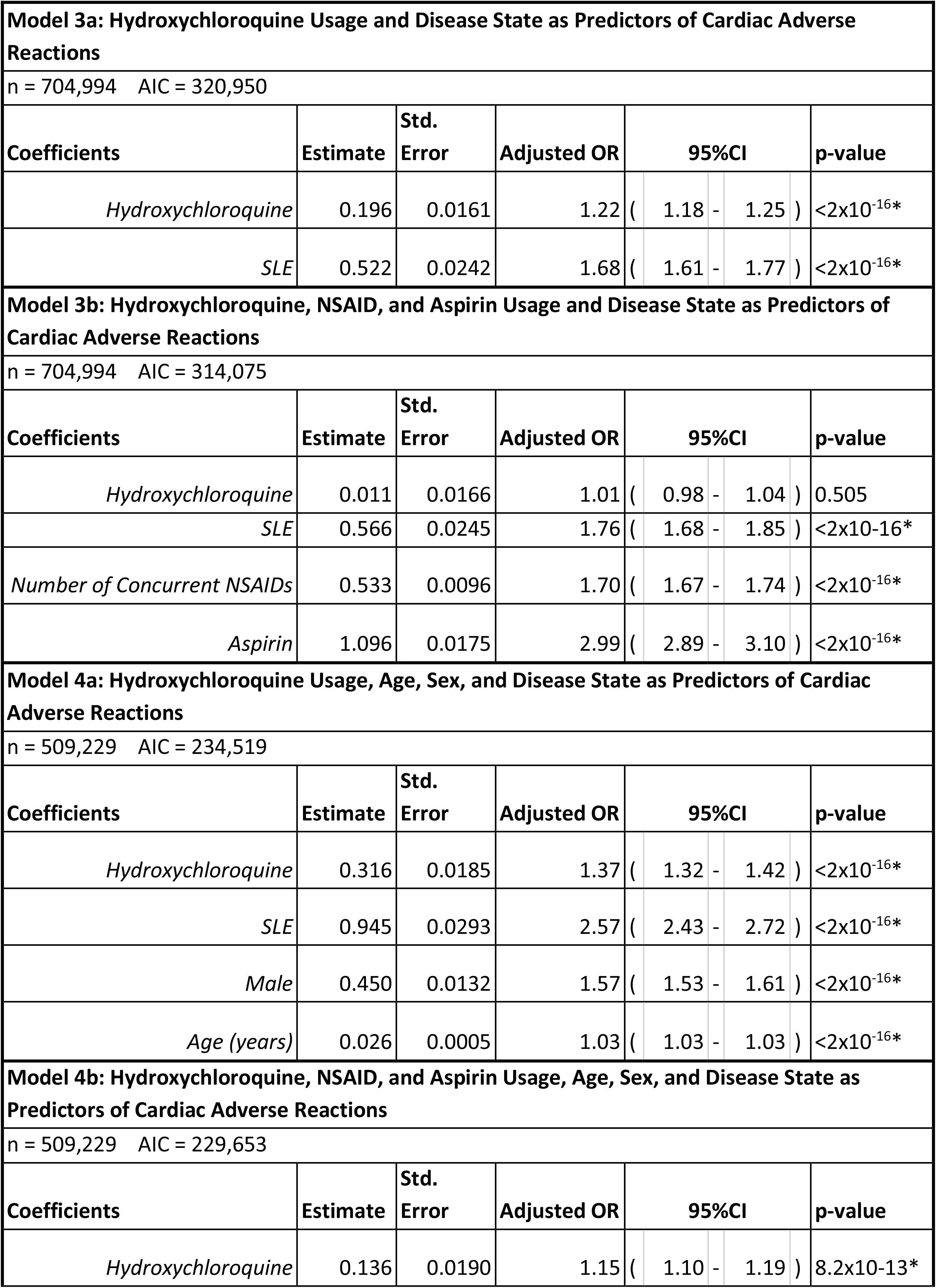
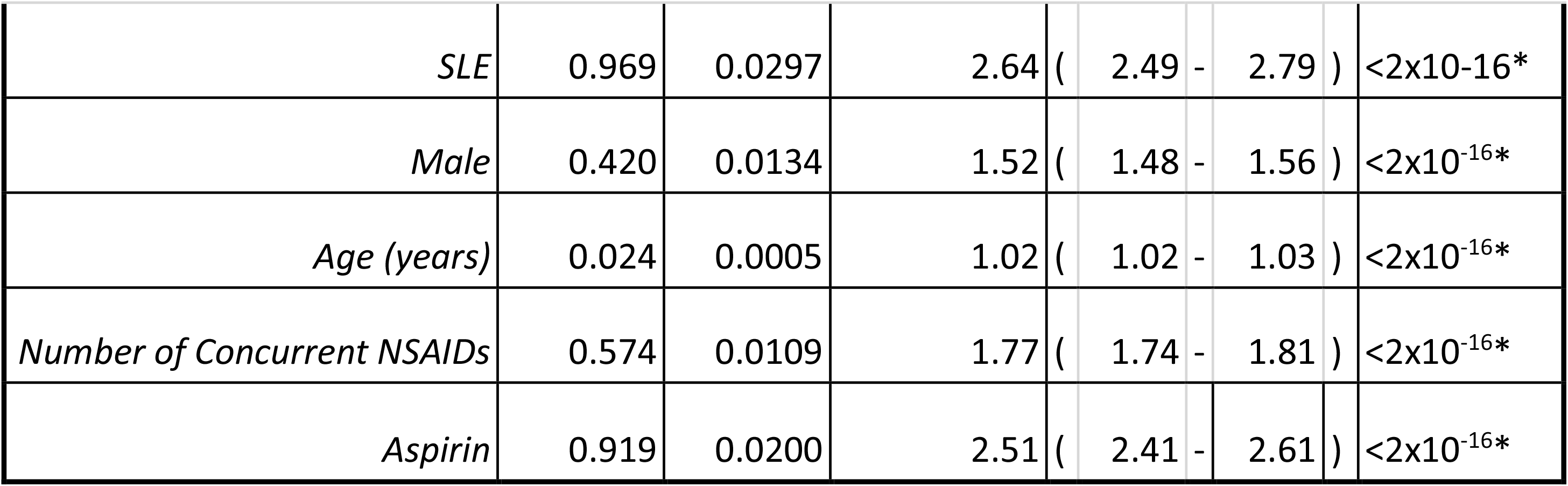
Logistic regression analysis of hydroxychloroquine AEs. * = coefficients with significant p-values

| Model 3a: Hydroxychloroquine Usage and Disease State as Predictors of Cardiac Adverse Reactions |  |  |  |  |  |  |
| --- | --- | --- | --- | --- | --- | --- |
| n = 704,994 AIC = 320,950 |  |  |  |  |  |  |
| Coefficients | Estimate | Std. Error | Adjusted OR | 95%CI |  |  |
| *Hydroxychloroquine* | 0.196 | 0.0161 | 1.22 ( | 1.18 | - 1.25 ) | <2x10<sup>-16</sup>* |
| *SLE* | 0.522 | 0.0242 | 1.68 ( | 1.61 | - 1.77 ) | <2x10<sup>-16</sup>* |
| Model 3b: Hydroxychloroquine, NSAID, and Aspirin Usage and Disease State as Predictors of Cardiac Adverse Reactions |  |  |  |  |  |  |
| n = 704,994 AIC = 314,075 |  |  |  |  |  |  |
| Coefficients | Estimate | Std. Error | Adjusted OR | 95%CI |  |  |
| *Hydroxychloroquine* | 0.011 | 0.0166 | 1.01 ( | 0.98 | - 1.04 ) | 0.505 |
| *SLE* | 0.566 | 0.0245 | 1.76 ( | 1.68 | - 1.85 ) | <2x10<sup>-16</sup>* |
| *Number of Concurrent NSAIDs* | 0.533 | 0.0096 | 1.70 ( | 1.67 | - 1.74 ) | <2x10<sup>-16</sup>* |
| *Aspirin* | 1.096 | 0.0175 | 2.99 ( | 2.89 | - 3.10 ) | <2x10<sup>-16</sup>* |
| Model 4a: Hydroxychloroquine Usage, Age, Sex, and Disease State as Predictors of Cardiac Adverse Reactions |  |  |  |  |  |  |
| n = 509,229 AIC = 234,519 |  |  |  |  |  |  |
| Coefficients | Estimate | Std. Error | Adjusted OR | 95%CI |  |  |
| *Hydroxychloroquine* | 0.316 | 0.0185 | 1.37 ( | 1.32 | - 1.42 ) | <2x10<sup>-16</sup>* |
| *SLE* | 0.945 | 0.0293 | 2.57 ( | 2.43 | - 2.72 ) | <2x10<sup>-16</sup>* |
| *Male* | 0.450 | 0.0132 | 1.57 ( | 1.53 | - 1.61 ) | <2x10<sup>-16</sup>* |
| *Age (years)* | 0.026 | 0.0005 | 1.03 ( | 1.03 | - 1.03 ) | <2x10<sup>-16</sup>* |
| Model 4b: Hydroxychloroquine, NSAID, and Aspirin Usage, Age, Sex, and Disease State as Predictors of Cardiac Adverse Reactions |  |  |  |  |  |  |
| n = 509,229 AIC = 229,653 |  |  |  |  |  |  |
| Coefficients | Estimate | Std. Error | Adjusted OR | 95%CI |  |  |
| *Hydroxychloroquine* | 0.136 | 0.0190 | 1.15 ( | 1.10 | - 1.19 ) | 8.2x10<sup>-13</sup>* |

|  |  |  |  |  |  |  |  |
| --- | --- | --- | --- | --- | --- | --- | --- |
| *SLE* | 0.969 | 0.0297 | 2.64 ( | 2.49 | - | 2.79 ) | <2x10<sup>-16</sup>* |
| *Male* | 0.420 | 0.0134 | 1.52 ( | 1.48 | - | 1.56 ) | <2x10<sup>-16</sup>* |
| *Age (years)* | 0.024 | 0.0005 | 1.02 ( | 1.02 | - | 1.03 ) | <2x10<sup>-16</sup>* |
| *Number of Concurrent NSAIDs* | 0.574 | 0.0109 | 1.77 ( | 1.74 | - | 1.81 ) | <2x10<sup>-16</sup>* |
| *Aspirin* | 0.919 | 0.0200 | 2.51 ( | 2.41 | - | 2.61 ) | <2x10<sup>-16</sup>* |

## Discussion

In our study we analyzed 702,274 FDA adverse event reports divided into CQ, HCQ and control cohorts to determine their association with cardiac AEs when taking into account other factors with increased cardiac risk. Although both drugs were significantly associated with increased cardiac AE risk, it was observed that this effect was substantially larger for CQ than HCQ (Fig. 1). Additionally, age, sex, concurrent NSAID use, and disease state were identified to contribute to the risk of cardiotoxicity with both therapeutics. After controlling for these factors, it was observed that the deleterious effects of CQ and HCQ on cardiac AE risk remained significant (Tables 2 and 3).

**Figure 1.**
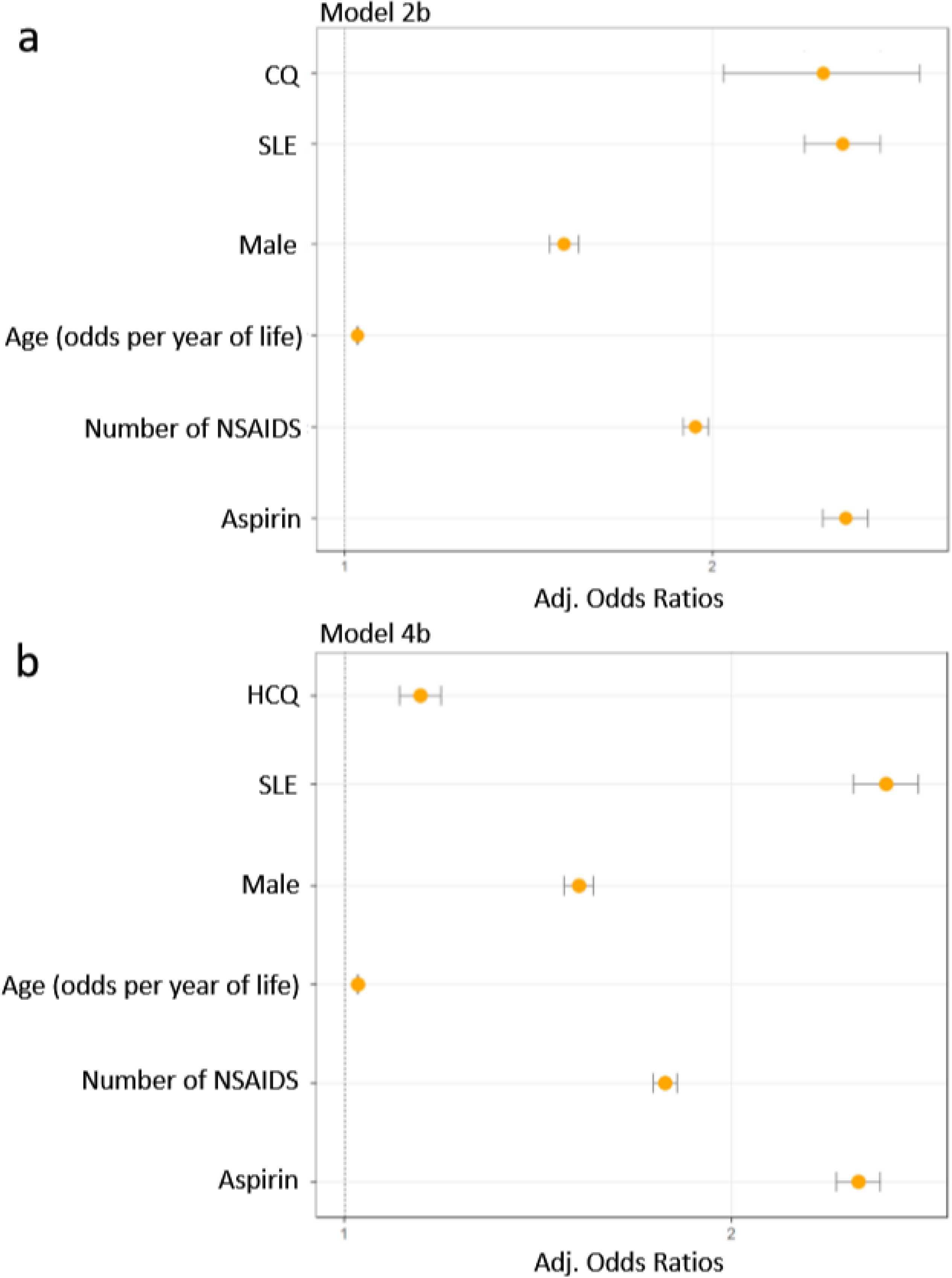
**Legend**. Adjusted Odds Ratios from Models 2b (n = 464,528) and 4b (n = 509,229) presented. (a) Chloroquine, NSAID, and aspirin usage, age, sex, and disease state as predictors of cardiac AEs (b) Hydroxychloroquine, NSAID, and aspirin usage, age, sex, and disease state as predictors of Cardiac AEs.

We expanded the current safety surveillance evidence by using a more comprehensive list of cardiac AEs (Supplementary Table S1), to avoid diluting the safety signal with many individual terms. Additionally, we performed multivariate analysis and identified several at-risk populations. The results of the multivariate binary logistic regression were validated by prior knowledge from literature: 1) *Sex*, when compared females, males are more often diagnosed with myocardial infarction, fatal coronary heart disease, among other cardiac diseases^38^; 2) *Age*, cardiovascular disease (CVD) has been shown to increase with age across multiple populations^38^; 3) *NSAIDs*, especially preferential and selective cyclooxygenase-2 inhibitors, increase the risk of CVD^39,40^.Two other essential factors were shown to be necessary to correct for, due to their association with cardiac side effects in the patient population used for our analysis and the current utilization of CQ and HCQ: 4) *SLE:* SLE patients have been shown to have a greater risk of cardiac AEs than patients with RA. This makes sense, as it is well known that CVD is one of the major complications and is one of the leading factors in mortality in patients living with SLE^41^. Additionally, the host inflammatory response seen in SLE may be similar to that in COVID^42,43^. 5) *Aspirin:* It may be expected for aspirin to present a protective effect, however the opposite effect was observed. Use of Aspirin in itself does not increase CVD risk, in fact it is mainly used for preventing cardiac events^44^. Aspirin use in our study population is likely heavily associated prior cardiac related medical history, that is not listed in FEARS, and could not be otherwise accounted for in the model. Aspirin increases the robustness of the model by correcting for prior treatment of cardiac disease or prevention in high risk patients. The regression model was instrumental to exclude this aspirin association from the quantification of the direct cardiac side effects of CQ and HCQ, which remained significant after adjustment (Adjusted Odds Ratios[95%CI]: 2.46[2.04-2.95] and 1.15[1.10-1.19] respectively).

### Generalizability of Results to COVID-19 Treatment with CQ and HCQ

This analysis revealed increased cardiac risk factors associated with CQ and HCQ that likely apply to a wide range of patients. Although our study was not performed on reports from Covid-19 patients, due to absence of such reports at the time of the study, the results may still be of value because of our mode of evaluation. Additionally, the cardiac complications and related mortality of SARS-CoV-2 patients^45^, are attributed to the inflammatory nature of the infection^42,43^, which is also seen in SLE pathophysiology ^46,47^. In fact, several case reports describe the heart related mortality being associated with inflammation of the myocardium ^48,49^. CQ and HCQ use in SLE and RA cohorts for the study was a beneficial coincidence, since they are also inflammatory conditions affecting the cardiovascular system that are also treated with CQ and HCQ^50,41^.

### Conclusion

In this study we observed increased risk of cardiac AEs in FAERS/AERS reports of CQ and HCQ with respect to other therapeutics used for RA and SLE. The association remained significant when demographic parameters and concurrent medications were accounted for in the analysis. It may be beneficial to closely monitor patients for cardiac complications. HCQ may be safer for use than CQ in patients at higher risk of cardiac complications. Although, when compared to CQ, HCQ use was associated with a lower risk of these events, the risk was still statistically significant. If and when available, alternative therapeutics may be safer to use for SARS-CoV-2 patients who are already at higher risk of cardiovascular complications due to age, pre-existing cardiovascular issues, concomitant medications and the SARS-CoV-2 infection itself.

## Data Availability

The study used over thirteen million AE reports available from the United States Food and Drug Administration Adverse Event Reporting System (FAERS) and its older version, Adverse Event Reporting System (AERS) data sets. At the time of the study the FAERS/AERS set contained reports from years 2000-2020, all available online at:
https://www.fda.gov/drugs/questions-and-answers-fdas-adverse-event-reporting-system-faers/fda-adverse-event-reporting-system-faers-latest-quarterly-data-files
Also made available here is the cleaned database "Cleaned_Data.csv" which can be used to run the supplied R code to reproduce the results.

## Study limitations

Due to the voluntary nature of the FAERS/AERS reports, actual population incidences of the adverse events cannot be derived. MedWatch reporting may also be biased by newsworthiness and legal variables. The safety surveillance data misses comprehensive medical records and medication history limiting the scope of the analysis. As with any association study, causality may not be derived from association, since the cases were not uniformly evaluated for causality by clinical specialists.

However, the postmarketing surveillance data analysis of over 700,000 reports provides population scale evidence which can be used to identify safety signals that might go unnoticed in small scale studies. While our approach with multivariate logistic regression controls for important biases in the data, such as sex, age, and concomitant drug use, some unaccounted-for factors may remain.

## Acknowledgements

We thank members of the Abagyan lab for support during this project. We also thank Da Shi for the contribution to processing the FAERS/AERS data sets.

## Author information

### Affiliations

Clinical Pharmacology and Therapeutics (CPT) Postdoctoral Training Program, University of California San Francisco, San Francisco, California, United States

Isaac V. Cohen

Skaggs School of Pharmacy and Pharmaceutical Sciences, University of California San Diego, La Jolla, California, United States

Tigran Makunts, Talar Moumedjian, Masara Issa & Ruben Abagyan

Oak Ridge Institute of Science and Education (ORISE), Clinical Pharmacology and Machine Learning fellowship at the Center for Drug Evaluation and Research, United States Food and Drug Administration, Silver Spring, Maryland, United States

Tigran Makunts

### Contributions

I.V.C and T.Ma performed the experiments. T.Ma, I.V.C, and R.A designed the study, I.V.C, T.Ma, T.M., M.A.I and R.A. drafted the manuscript and reviewed the final version. R.A. processed the data. I.V.C and T.Ma contributed equally to this work.

## Ethics declarations

Authors declare no conflict of financial or non-financial interest.

